# Prevalence of falls in the last weeks of life and relationship between falls, independence, and quality of dying: A secondary analysis of a large prospective cohort study

**DOI:** 10.1101/2024.02.12.24302685

**Authors:** Hiroyuki Otani, Junichi Shimoinaba, Hideyuki Kashiwagi, Tatsuya Morita, Isseki Maeda, Naosuke Yokomichi, Jun Hamano, Takashi Yamaguchi, Masanori Mori

**Affiliations:** Department of Palliative and Supportive Care, St. Mary’s Hospital, 422 Tsubukuhonmachi, Kurume city, Fukuoka 830-8543, Japan; Department of Palliative Care Team and Palliative and Supportive Care, National Hospital Organization Kyushu Cancer Center, 3-1-1 Notame, Mitami-ku, Fukuoka 811-1395, Japan; Department of Palliative Care, Eiko Hospital, 3-8-15 Beppu Nishi, Shimecho, Kasuya-gun, Fukuoka 811-2232, Japan; Department of Transitional and Palliative Care, Iizuka Hospital, 3-83 Yoshiocho, Iizuka City, Fukuoka 820-0018, Japan; Palliative and Supportive Care Division, Seirei Mikatahara General Hospital, 3453 Mikatahara-cho, Kita-ku, Hamamatsu city, Shizuoka 433-8558, Japan; Department of Palliative Care, Senri-chuo Hospital, 1-4-3 Shinsenrihigashi-machi, Toyonaka city, Osaka 560-0082, Japan; Department of Palliative and Supportive Care, Institute of Medicine, University of Tsukuba 1-1-1 Tennoudai, Tsukuba, Ibaraki 305-8575, Japan; Department of Palliative Medicine, Kobe University Graduate School of Medicine, 7-5-1 Kusunoki-cho, Chuo-ku, Kobe 650-0017, Japan

**Keywords:** Falls, Terminally Ill Cancer Patients, independence, quality of dying, fall prevention

## Abstract

**Objective:** To determine the frequency of falls and their serious complications in palliative care units (PCUs), as well as explore the complex interplay between falls, independence, and quality of dying.

**Methods:** A secondary analysis of a large prospective cohort study of 23 PCUs in Japan was conducted from January 2017 to June 2018. Palliative care specialist physicians recorded whether patients experienced falls, serious complications from falls, activities that led to falls, independence (workability in the last days and use of indwelling urinary catheter), and Good Death Scale.

**Results:** Of the 1,633 patients evaluated, 9.2% (95% Confidence interval [95% CI 7.8 to 11]) experienced falls within 30 days prior to death. The patients who fell were mostly men, had eastern cooperative oncology group performance status 3 on admission, a longer estimated prognosis on admission, and delirium during hospitalization. Serious falls causing fractures or intracranial hemorrhages were rare (0.3% [95% CI 0.038 to 0.57]). The most common reason for falls was the need to use the toilet. The Good Death Scale and indwelling urinary catheter use were not significantly associated with falls.

**Conclusion:** Falls occur in approximately 10% of patients in PCUs, but serious complications are rare. The relationship between falls, independence, and quality of dying is complex; that is, a fall may not be necessarily bad, if it is the result of respect for the patient’s independence. Healthcare providers need to consider fall prevention while supporting patients’ desire to move on their own to maintain independence.

**WHAT IS ALREADY KNOWN ON THIS TOPIC:**

- Falls are a major healthcare concern because of their potential to cause physical harm, emotional distress, and increased healthcare costs.
- Although many studies have investigated falls in acute care settings and the elderly population, there is a lack of literature specifically focusing on falls in the unique context of palliative care units.

**WHAT THIS STUDY ADDS:**

- Falls occur in only approximately 10% of patients, and only five cases (0.3%) of serious events were due to falls in palliative care units (PCUs).
- Independence and quality of dying are not significantly compromised by falls.

**HOW THIS STUDY MIGHT AFFECT RESEARCH, PRACTICE OR POLICY:**

- Although fall prevention is considered a priority, healthcare providers should support patients’ desires to move on their own to maintain independence.
- It may be possible to maintain independence and quality of dying even for patients who have fallen.

## 1 INTRODUCTION

Falls are a major healthcare concern because of their potential to cause physical harm, emotional distress, and increased healthcare costs. Although falls are often perceived as a problem that is closely associated with acute care settings, they can also have a significant effect on patients in palliative care units (PCUs) [1,2]. These patients are usually in the final stages of their illness and may have reduced mobility and strength, increasing their dependence on care and support. Falls in this vulnerable stage can lead to fractures, bleeding, and other serious complications [3–5], increasing the burden faced by terminally ill patients and causing pain and suffering beyond the physical realm.

However, the consequences of falls during the last weeks of life are not limited to physical injuries. This resonates throughout the broader narrative of patient autonomy and quality of dying. For patients receiving palliative care, the ability to move around, contribute to their care, and make appropriate choices is critical. Falls threaten their autonomy and violate the cherished sense of dignity they hope to maintain. Loss of independence, even later in life, can be a serious emotional experience for patients, increasing their vulnerability and reducing their overall quality of life. Even a single fall can lead to excessive anxiety and self-doubt about moving, and even loss of motivation to live owing to the fear of falling again [6–10]. Although the topics of falls among terminally ill cancer patients and terminally ill patients’ sense of autonomy and quality of dying have been investigated separately [11–12], research on the interplay between both variables is limited. Moreover, research on the relationship between falls and the quality of death has only been conducted from a safety perspective [13].

Understanding the frequency of falls in PCUs is critical for several reasons. First, it provides important data for healthcare providers so that risk factors specific to this population can be identified and addressed. Second, it helps to develop targeted interventions to reduce the risk of falls and promote patient safety. Third, it will help improve the overall quality of care provided in palliative care settings, with the overarching goal of enhancing the patient’s end-of-life experience.

Although many studies have investigated falls in acute care settings and the elderly population, there is a lack of literature specifically focusing on falls in the unique context of PCUs.

This study primarily aimed to determine the frequency of falls and serious falls in PCUs. The secondary aim was to explore the complex interplay between independence and the quality of dying in the presence of falls.

## 2 MATERIALS AND METHODS

### 2.1 Study design and subjects

This study involved a secondary analysis of a large prospective cohort study conducted between January 2017 and June 2018 to investigate the dying process of patients with advanced cancer in 23 PCUs in Japan [14]. Consecutive eligible patients were enrolled if they had been newly referred to the participating PCUs during the study period, and all enrolled patients were observed until their death or six months after enrollment. All 23 institutions were asked to collect data consecutively, up to a designated number of patients of 50, 60, 70, 80, 100, 150, and 250, according to the size of the institution, without depending on high-volume centers. This study’s inclusion criteria were as follows: (1) adult patients (18 years of age or older), (2) patients diagnosed with locally advanced or metastatic cancer (including hematological neoplasms), and (3) patients admitted to PCUs. Patients who were planned to be discharged within one week or those who did not want to be enrolled were excluded. This study upheld ethical standards per the Helsinki Declaration and Japanese Ministry of Health guidelines for research involving human subjects, with written consent waived for this noninvasive observational study. It was also approved by the institutional review boards at all participating sites.

### 2.2 Measurements

#### 2.2.1 Demographic and clinical characteristics

The primary physician most involved with the patient obtained prospective data at the time of death from daily clinical practice, including eastern cooperative oncology group (ECOG) performance status on admission, estimated prognosis on admission, the presence or absence of delirium during admission, and opioid use during admission. Demographic data, including age, sex, primary cancer site, length of hospital stay days, and presence or absence of bone and central nervous system metastases on admission [3], were obtained from the patients’ medical charts.

#### 2.2.2 Falls

The primary physician most involved with the patient obtained prospective data from daily clinical practice, including experiences of falls, reasons for behavior-led falls according to previous studies (i.e., moving to use the toilet to defecate, picking up things in the room, washing one’s face, among others; [15–16]), and serious complications related to falls up to 30 days prior to death. Serious complications were defined as fractures and intracranial hemorrhage according to previous studies [1,3,5]. The observation period was set at 30 days before death rather than during the entire hospital stay, because our research interest was on falls that occurred in the last days, and most patients died within 30 days from admission.

#### 2.2.3 Good Death Scale

In this study, we adopted the good death scale (GDS) as an indicator of quality of dying. The GDS is a provider assessment scale for the quality of dying in patients with terminal illnesses and cancer. The linguistic validity of the Japanese version has been confirmed [17–21]. The GDS comprises five domains: the awareness that one is dying (0, complete ignorance; 1, ignorance; 2, partial awareness; 3, complete awareness), peaceful acceptance of death (0, complete unacceptance; 1, unacceptance; 2, acceptance; 3, complete acceptance), honoring of the patient’s wishes (0, no reference to the will of either the patient or the family; 1, following the family’s will alone, 2, following the patient’s will alone; 3, following the will of both), death timing (0, no preparation; 1, family alone had prepared; 2, patient alone had prepared; 3, both had prepared well), and the degree of physical comfort three□days before death (0, a lot of suffering; 1, suffering; 2, a little suffering; 3, no suffering). The GDS was evaluated by the attending physician after each patient’s death.

#### 2.2.4. Variables related to independence

We recorded data on patients’ use of indwelling urinary catheter during admission and walkability until three or seven days before death as indicators of independence.

### 2.3 Statistical analyses

We analyzed patients who experienced falls one month prior to death as the primary endpoint. The baseline demographic data were summarized using descriptive statistics. First, we calculated the prevalence of falls in all the patients. Additionally, we conducted a subgroup analysis for the patients with a performance status of 3 or less (i.e., excluding the patients with a performance status of 4), on the assumption that bedridden patients had no theoretical risk of falls but some patients with a performance status of 4 might recover their physical function after admission. Second, we compared the patient backgrounds between the patients who did and did not experience falls. Univariate analyses were performed using the Student’s t-test or chi-square test, as appropriate. Multiple logistic regression analyses were then performed using a forward elimination procedure. Third, we calculated the prevalence of serious complications from falls in all the patients and the patients with a performance status of 3 or less. Fourth, we determined the clinical factors associated with falls and serious complications from falls for each patient. Fifth, we calculated the percentage of activities that led to falls in the patients who experienced them. Finally, we compared the data on independence (i.e., use of indwelling urinary catheter during admission and walkability until three or seven days before death) and quality of dying measured by the GDS between the patients who did and did not fall using the Student’s t-test or chi-square test, as appropriate. Subgroup analyses were also performed in this comparison. Statistical significance was set at P <.05. All analyses were performed using SPSS version 28.0.1 (IBM Corp., Armonk, NY, USA).

## 3 RESULTS

In total, 2591 patients with cancer were admitted to the participating 23 PCUs in Japan between January 2017 and June 2018. Of those, 1971 patients were assessed for eligibility. After excluding 45 patients (scheduled to be discharged within a week: 42 patients, patient or family declined to participate: 3 patients), we finally enrolled 1926 patients and analyzed the data of 1896 patients. Of the patients analyzed, 263 were excluded because they were discharged alive. Thus, the total number of patients evaluated in this study was 1633 (Figure 1). The patient characteristics are summarized in Table 1. The median age was 74 years; approximately 40% experienced delirium during hospitalization, and approximately 80% used opioids during hospitalization; moreover, the median length of hospital stay days was 16 days. The number of the patients with performance status of 4 was 860 (52.6%), and thus the remaining 773 patients had performance status of 1 to 3.

**Figure 1.**
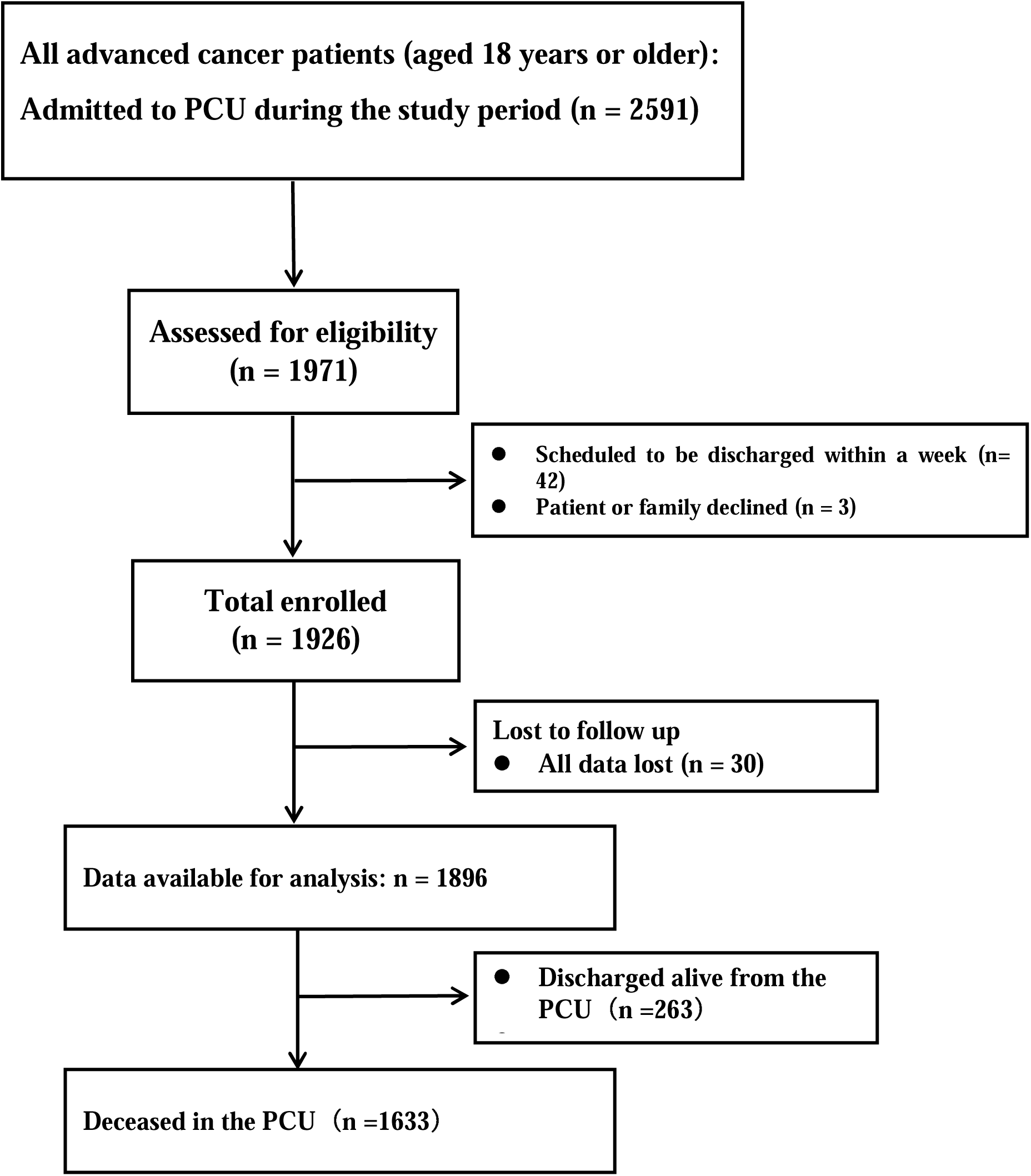
Flow chart per STROBE

**Table 1.**
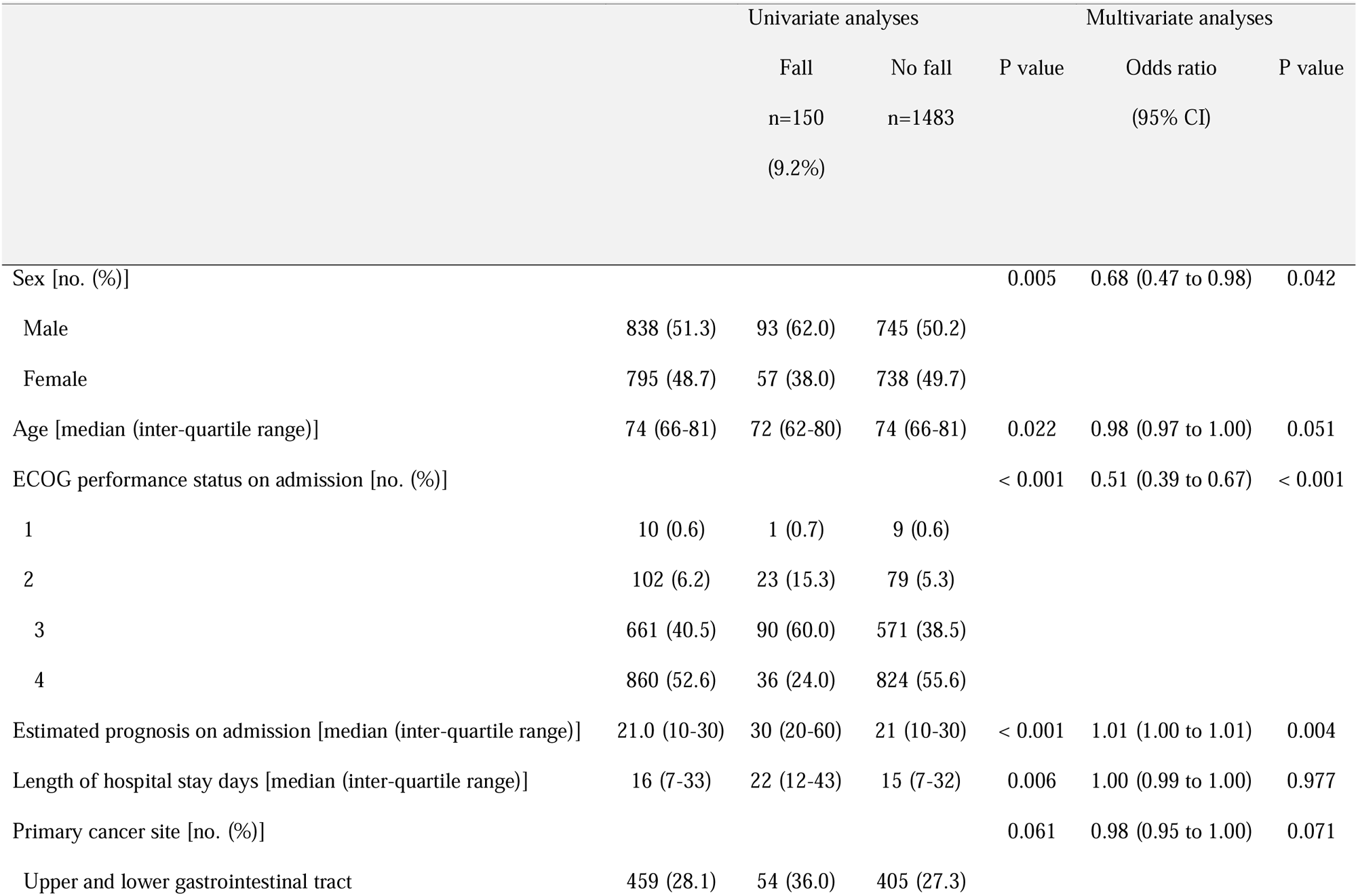
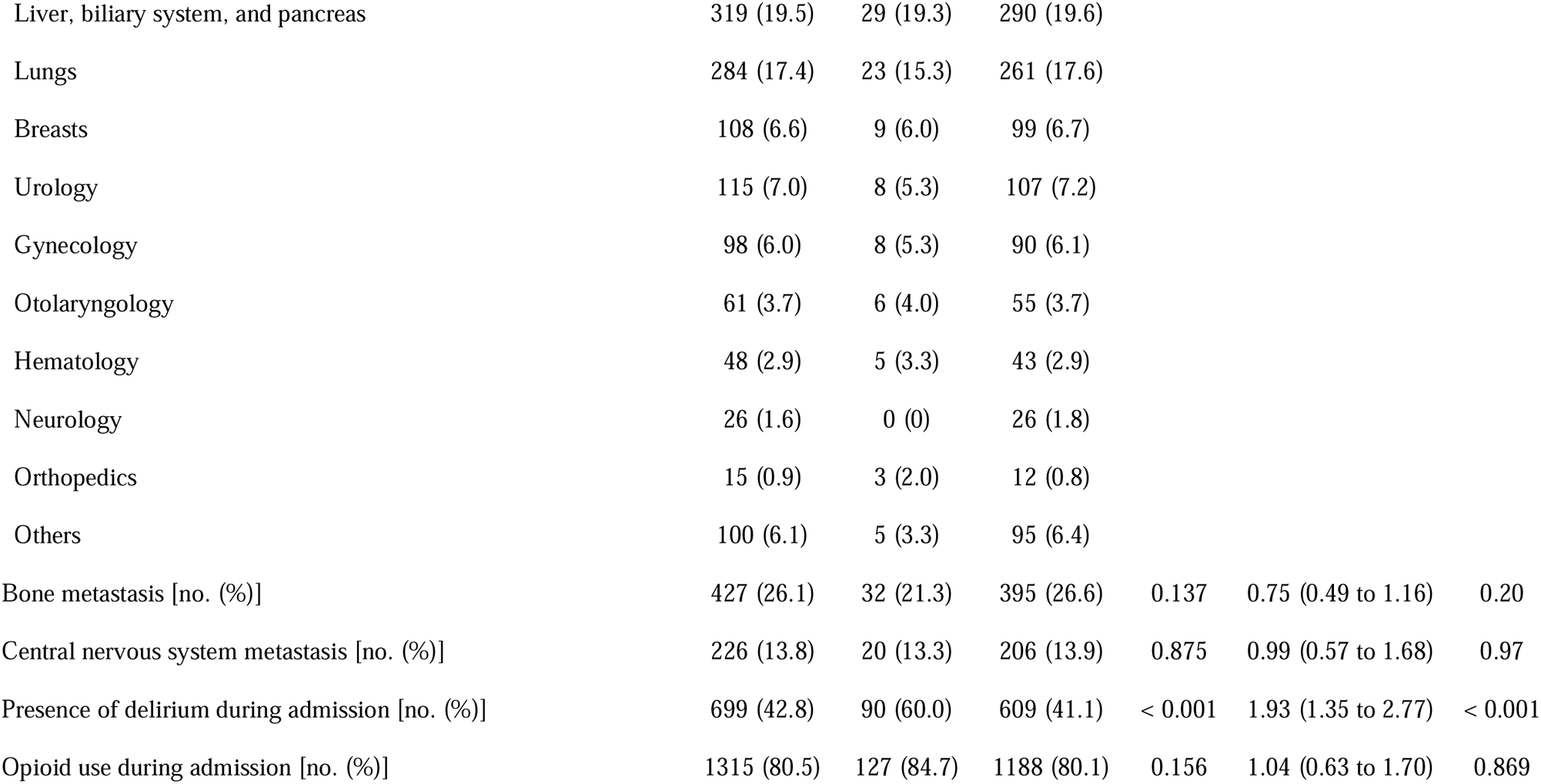
Patient characteristics and comparisons of the patients who did and did not experience falls (n=1633)

### 3.1 Prevalence of fall

Of the total number of patients evaluated, 150 (9.2% [95% CI 7.8 to 11]) fell within 30 days prior to death; limiting the sample to patients with a performance status of 3 or less on admission, 114 (14.8% [95% CI 12 to 17]) fell within 30 days prior to death.

Patients who fell were more likely to be male, younger, had the ECOG performance status 3 on admission, had a longer length of hospital stay days and longer estimated prognosis by the primary physician on admission, and had more delirium during hospitalization (Table 1). The multiple logistic regression analysis revealed that independent determinants of falls were being male, had the ECOG performance status 3 on admission, longer estimated prognosis by the primary physician on admission, and the presence of delirium during hospitalization.

### 3.2 Serious complication of fall

There were five cases of serious events due to falls, three with fractures, and two with intracranial hemorrhages (Table 2). This figure corresponds to 0.31% [95% CI 0.038 to 0.57] in all patients, and 0.52% [95% CI 0.012 to 1.0] in patients with a performance status of 3 or less (Table 2). Table 3 describes the factors potentially associated with falls, fracture, and hemorrhage in each patient. Both patients with intracranial hemorrhages were elderly (79 and 91 years old); one younger patient with a fracture had bone metastases, whereas two older patients with fractures did not (81 and 83 years old). All patients received opioid medications, and 3 of 5 had an episode of delirium and received antipsychotics. None of the two patients who developed intracranial hemorrhage had risk factors of hemorrhage (i.e., severe liver dysfunction, alcohol abuse, or low platelet count).

**Table 2.** Frequency of serious events due to falls.

| Table 2. Frequency of serious events due to falls |  |  |  |  |  |  |  |  |  |  |
| --- | --- | --- | --- | --- | --- | --- | --- | --- | --- | --- |
| Total (n=1633) |  |  |  |  |  | Patients with ECOG performance statuses of 0–3 on admission (n=773) |  |  |  |  |
|  | All patients (n=1633) |  |  | Patients who experienced falls (n=150) |  | All patients (n=773) |  |  | Patients who experienced falls (n=114) |  |
|  | N | % | 95%CI | % | 95%CI | n | % | 95%CI | % | 95%CI |
| Any serious complications | 5 | 0.31 | 0.038 to 0.57 | 3.3 | 0.46 to 6.2 | 4 | 0.52 | 0.012 to 1.0 | 3.5 | 0.13 to 6.9 |
| Fracture due to fall | 3 | 0.18 | 0 to 0.39 | 2.0 | 0 to 4.2 | 3 | 0.39 | 0 to 0.83 | 2.6 | 0 to 5.6 |
| Intracranial hemorrhage due to fall | 2 | 0.12 | 0 to 0.29 | 1.3 | 0 to 3.2 | 1 | 0.13 | 0 to 0.38 | 0.88 | 0 to 2.6 |

**Table 3.** Patients who experienced serious complications related to falls.

|  |  |  |  |  |  |
| --- | --- | --- | --- | --- | --- |
|  | Case 1 | Case 2 | Case 3 | Case 4 | Case 5 |
| **Serious events** | Fracture | Fracture | Fracture | Intracranial hemorrhage | Intracranial hemorrhage |
| **Demographic data** |  |  |  |  |  |
| Sex | Female | Female | Female | Male | Male |
| Age | 83 | 81 | 60 | 91 | 79 |
| Primary cancer site | Upper and lower<br>gastrointestinal tract | Hematology | Upper and lower<br>gastrointestinal tract | Upper and lower<br>gastrointestinal tract | Upper and lower<br>gastrointestinal tract |
| Length of hospital stay (days) | 12 | 24 | 23 | 7 | 23 |
| **Factors potentially associated with fall** |  |  |  |  |  |
| Central nervous system metastasis | None | None | None | None | None |
| Presence of delirium during admission | None | Present | Present | None | Present |
| Antipsychotic use during admission | None | Present | Present | None | Present |
| Opioid use during admission | Present | Present | Present | Present | Present |
| Daily opioid dose (oral morphine equivalent) at death | 30 | 30 | 100 | 240 | 20 |
| **Factors potentially associated with fracture** |  |  |  |  |  |
| Bone metastasis | None | None | Present | Present | None |
| **Factors potentially associated with hemorrhage** |  |  |  |  |  |
| Severe liver dysfunction on admission | None | None | None | None | None |
| Alcohol abuse disorder on admission | None | None | None | None | None |
| Platelet count on admission | $56.5 \times 10^4/\mu\text{L}$ | $31.7 \times 10^4/\mu\text{L}$ | $22.5 \times 10^4/\mu\text{L}$ | $21.9 \times 10^4/\mu\text{L}$ | $40.5 \times 10^4/\mu\text{L}$ |

### 3.3 Activities that led to falls

The behaviors that led to falls were: going to the toilet to defecate (64.7%; 97/150), picking up things in the room (12.7%; 19/150), and washing their face and hands (1.3%, 2/150) (Table 4).

**Table 4.** Behavior that led to falls (n=150) [no. (%)]

|  |  |
| --- | --- |
| Moving to the toilet for elimination | 97 (64.7%) |
| Picking up things in the room | 19 (12.7%) |
| Washing one's face | 2 (1.3%) |
| Unknown | 32 (21.3%) |

### 3.4 Variables related to independence and quality of dying

Patients who fell were more likely to be able to walk in the last days of life (Table 5). The GDS results and indwelling urinary catheter use were not significantly associated with falls.

**Table 5.**
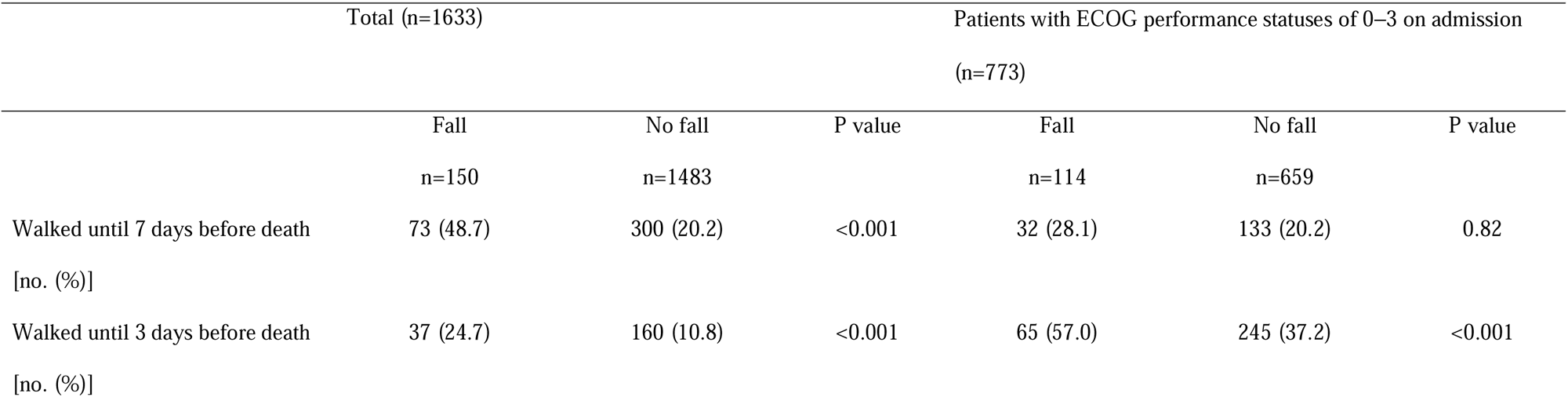
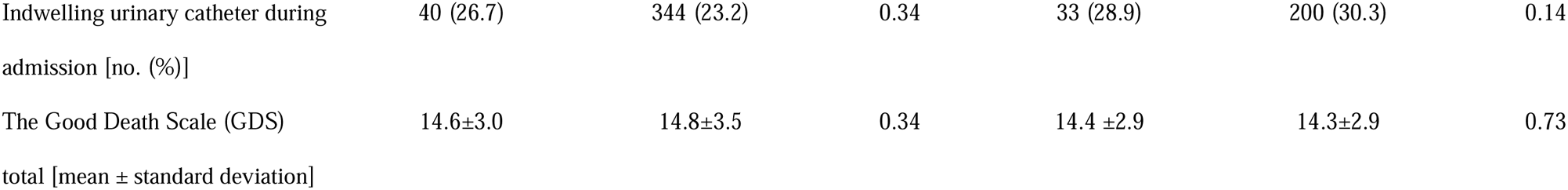
Variables related to independence and quality of dying.

|  | Total (n=1633) |  |  | Patients with ECOG performance statuses of 0–3 on admission (n=773) |  |  |
| --- | --- | --- | --- | --- | --- | --- |
|  | Fall<br>n=150 | No fall<br>n=1483 | P value | Fall<br>n=114 | No fall<br>n=659 | P value |
| Walked until 7 days before death<br>[no. (%)] | 73 (48.7) | 300 (20.2) | <0.001 | 32 (28.1) | 133 (20.2) | 0.82 |
| Walked until 3 days before death<br>[no. (%)] | 37 (24.7) | 160 (10.8) | <0.001 | 65 (57.0) | 245 (37.2) | <0.001 |
| Indwelling urinary catheter during admission [no. (%)] | 40 (26.7) | 344 (23.2) | 0.34 | 33 (28.9) | 200 (30.3) | 0.14 |
| The Good Death Scale (GDS) total [mean ± standard deviation] | 14.6±3.0 | 14.8±3.5 | 0.34 | 14.4 ±2.9 | 14.3±2.9 | 0.73 |

## 4 DISCUSSIONS

This large, multicenter, prospective, observational study revealed the frequency of falls and serious falls in PCUs. An important finding was the frequency and causes of falls and the complex interplay between independence and quality of dying in the presence of falls in PCUs.

### 4.1 Prevalence of falls in palliative care settings

In this study, 150 patients (9.2%) fell within 30 days prior to death in PCUs. Limiting the sample to patients with a performance status of 3 or less, this figure slightly increased to 14.7%. These findings are consistent with those of previous studies demonstrating that 11%–18% of patients in PCUs experienced falls [1,2,22–25]. Given that the reported fall rate for patients with advanced cancer is approximately 50% (3,26), patients in PCUs were found to have a relatively lower rate. This may be because of the more serious physical condition of the PCU patients and of the medical teams supporting patients in the palliative care ward. This study confirmed that approximately 10%−20% of patients experienced falls in palliative care units.

In addition, this study explored the factors associated with falls. Patients who fell were more likely to have the following characteristics: male, an ECOG performance status of 3 on admission, longer estimated prognosis by the primary physician on admission, and presence of delirium during hospitalization. Patients who fell were more likely to have more delirium during hospitalization. Previous studies [4,22] have shown that delirium is more likely to result in falls. Delirium increases the risk of falls because it can manifest as rapid fluctuations in alertness, attention, and behavior. Moreover, the neuroleptics/antipsychotics often used to treat it may increase the risk of falls [4,27]. In addition, in this study, men were more likely to fall. Previous studies also found that men were more likely to fall [22,25]. However, one study concluded that sex is not statistically significant in relation to fall risk (4). Therefore, it seems best to use these risks as a supplement to clinical judgment to address the continually shifting symptomatology of patients in terminal decline because changes in unstable situations are frequent in hospice care, such as the ECOG performance status 3 on admission, which has a significantly higher risk of falling in this study.

### 4.2 Prevalence of serious cases in patients who experienced falls

There were five cases (0.31% of all patients and 0.52% of patients with performance statuses of 1−3 on admission) of serious events due to falls, three with fractures, and two with intracranial hemorrhage. These figures in the current study are lower than those reported in previous studies on serious injuries (fractures) requiring severe medical treatment in a palliative care setting (1.6%–4.1%) [1,3], in an acute care setting (6.3%), and in situations with fall prevention measures (4.1%) [5]. This may indicate that the five (0.3%) serious events might not have been caused by the palliative care setting. In addition, both patients with intracranial hemorrhage were older adults, and had no risk factors of hemorrhage. Further, a younger patient with a fracture had bone metastases and two older patients had no bone metastases. This finding suggests that intracranial hemorrhage observed is related to the cases more likely to occur in older patients (e.g., subdural hematoma). Moreover, fracture is related to bone metastases in the younger population and osteoporosis in older patients. Serious events due to falls are extremely rare but they do occur, and not only in the palliative care setting. Although prevention is theoretically important because of the potentially devastating physical and emotional consequences, special precautions simply based on the setting may be unnecessary because they do not necessarily occur frequently in palliative care settings.

### 4.3 Relationship between falls, independence, and the quality of dying

The most common behavioral reason for falling in terminal cancer patients was “moving to the toilet to defecate.” This means that casual guidance and measures for the patient to use the toilet may reduce the frequency of falls and preserve the dignity of terminally ill cancer patients [15–16,28–30]. In particular, humiliation and shame over the failure to use the toilet have a significant bearing on the dignity of terminally ill cancer patients [16,31], and support for toileting is an issue that cannot be separated from the association between falls and dignity [32]. However, five reasons for resisting asking for assistance to go to the toilet are: they did not want someone to help them with personal activities; if elimination was an emergency, it would be difficult to wait for help to arrive; they thought they could move to the toilet on their own without falling; the nurses were too busy to help; and they did not want to ask for assistance from a nurse who they thought was not compassionate. Even providing elimination assistance involved complex factors [16].

We considered indwelling urinary catheter use and walkability in the last three or seven days of life as indicators of independence, and the GDS as an indicator of quality of dying. These were not associated with falls. Patients who fell were more likely to be able to walk in the last three or seven days of life (Table 5). As noted above, the most common reason for falls is defecation, but neither more nor fewer urinary catheters are used by those who fall. These results represent a significant effort by healthcare providers to pay more attention to the spiritual pain of patients in hospice settings. In addition, many people can walk until the end of their lives, with or without falls, probably as a result of healthcare providers supporting them to walk on their own. Furthermore, falling was not associated with the GDS scores. In previous studies, patients who fell were more likely to have a “greater fear of losing independence,” “avoid asking for help,” and “feel uneasy about asking for help” [4,6]. Nevertheless, in this study involving healthcare providers in a hospice setting, the GDS was not influenced by the presence or absence of falls, indicating that although fall prevention is considered a priority, healthcare providers should support patients’ desires to move on their own to maintain independence, and it may be possible to maintain independence and quality of dying even for patients who have fallen.

### 4.4 Strengths and limitations

This study’s strengths include a large sample of patients, the high participant response rate, multicenter design, and use of validated measurement tools. However, there are some limitations. First, the same palliative physician performed all observations for each patient. Therefore, observer bias and underreported bias must be considered. However, it is unlikely that the physician would have overlooked a fall because he or she is required to medically examine the patient at the time of the fall. Second, not all possible causes of falls are considered, such as past fall history or the effects of medications [3]. Third, we considered indwelling urinary catheter use during admission and walkability in the last three or seven days of life as indicators of independence and the GDS as an indicator of quality of dying. Our study involved secondary analyses of existing data, and we recognize that these factors alone cannot measure independence and the quality of dying. Similarly, we could not obtain information on the exact time when the falls occurred from the data. Therefore, some variables may have an inverse time relationship to the outcome variable (falls). Finally, we included only patients with cancer admitted to inpatient hospices/PCUs in Japan, where interprofessional care is provided for dying patients.

## 5. CONCLUSION

Approximately 10% of patients in PCUs experience inpatient falls within 30 days before death. Furthermore, 0.3% of the patients experienced severe cases. The main behavior that led to falls was going to the toilet to defecate; patients who fell were more likely to be able to walk in the last days of life, and neither the GDS results nor indwelling urinary catheter use was associated with experience of falls. Serious complications from falls were rare in palliative care settings, and relationship between falls, independence, and quality of dying was complex. A fall is not necessarily bad, if it is the result of respect for the patient’s independence to use the toilet by themselves. Healthcare providers need to consider fall prevention while paying attention to supporting patients’ desire to move on their own, with or without falls, to maintain independence, and further research is required in this domain.

## Author contributions

**Study concept and design:** Hiroyuki Otani, Junichi Shimoinaba, Hideyuki Kashiwagi, Tatsuya Morita, Isseki Maeda, Naosuke Yokomichi, Jun Hamano, Takashi Yamaguchi, Masanori Mori.

**Collection and/or assembly of data:** Hiroyuki Otani, Junichi Shimoinaba, Hideyuki Kashiwagi, Tatsuya Morita, Isseki Maeda, Naosuke Yokomichi, Jun Hamano, Takashi Yamaguchi, Masanori Mori.

**Statistical analysis:** Hiroyuki Otani, Junichi Shimoinaba, Hideyuki Kashiwagi, Tatsuya Morita, Isseki Maeda, Naosuke Yokomichi, Jun Hamano, Takashi Yamaguchi, Masanori Mori.

**Data analysis and interpretation:** Hiroyuki Otani, Junichi Shimoinaba, Hideyuki Kashiwagi, Tatsuya Morita, Isseki Maeda, Naosuke Yokomichi, Jun Hamano, Takashi Yamaguchi, Masanori Mori.

**Drafting of the manuscript:** Hiroyuki Otani, Junichi Shimoinaba, Hideyuki Kashiwagi, Tatsuya Morita, Isseki Maeda, Naosuke Yokomichi, Jun Hamano, Takashi Yamaguchi, Masanori Mori.

**Final approval of the manuscript:** Hiroyuki Otani, Junichi Shimoinaba, Hideyuki Kashiwagi, Tatsuya Morita, Isseki Maeda, Naosuke Yokomichi, Jun Hamano, Takashi Yamaguchi, Masanori Mori.

## Institutional review board statement

This study was approved by the local institutional review boards of all participating institutions, including the research ethics board of Seirei Mikatahara General Hospital research ethics board (no. 16-22). This study complied with the ethical standards of the Declaration of Helsinki and was performed following the ethical guidelines for epidemiological research presented by the Ministry of Health, Labour, and Welfare in Japan.

## Informed consent statement

We employed an opt-out method rather than acquiring written or oral informed consent because we were not required to obtain individual informed consent from participants in a non-invasive observational trial by Japanese law.

## Data availability

The datasets generated and analyzed during this study are not publicly available because sharing is not explicitly covered by patient consent.

